# The Concept of Stroma AReactive Invasion Front Areas (SARIFA) as a New Prognostic Biomarker for Lipid-driven Cancers Holds True in Pancreatic Ductal Adenocarcinoma

**DOI:** 10.1101/2024.01.22.24301622

**Authors:** Przemyslaw Grochowski, Bianca Grosser, Florian Sommer, Andreas Probst, Johanna Waidhauser, Gerhard Schenkirsch, Nic G. Reitsam, Bruno Märkl

## Abstract

**Background:** Pancreatic ductal adenocarcinoma (PDAC) is a ‘difficult-to-treat’ entity. To forecast its prognosis, we introduced a new biomarker, SARIFA (stroma areactive invasion front areas), which are an area at the tumour invasion front lacking desmoplastic stroma reaction upon malignant invasion in the surrounding tissue, leading to direct contact between tumour cells and adipocytes. SARIFA showed its significance in gastric and colorectal carcinoma, revealing lipid metabolism alternations that promote tumour progression.

**Methods:** We reviewed the SARIFA status of 174 PDAC cases on all available H&E-stained tumour slides from archival Whipple-resection specimens. SARIFA positivity was defined as SARIFA detection in at least 66% of the available slides. To investigate alterations in tumour metabolism and microenvironment, we performed immunohistochemical staining for FABP4, CD36 and CD68. To verify and quantify a supposed delipidation of adipocytes, adipose tissue was digitally morphometrised.

**Results:** In total, 54 cases (31%) were classified as SARIFA positive and 120 (69%) as SARIFA negative. Patients with SARIFA-positive PDAC showed a significantly worse overall survival compared with SARIFA-negative cases (median overall survival: 9.9 months vs. 18.0 months, HR: 1.558 (1.081–2.247), 95% CI, p = 0.018), which was independent from other prognostic markers (p = 0.014). At the invasion front of SARIFA-positive PDAC, we observed significantly higher expression of FABP4 (p<0.0001) and higher concentrations of CD68^+^ macrophages (p=0.031) related to a higher risk of tumour progression. CD36 staining showed no significant expression differences. The adipocyte areas at the invasion front were significantly smaller, with mean values of 4021 ± 1058 µm^2^ and 1812 ± 1008 µm^2^ for the SARIFA-positive and -negative cases, respectively. The area differences between the SARIFA-positive invasion front area and the other three parameters were highly significant (p < 0.001)

**Conclusions:** SARIFA is a promising prognostic biomarker for PDAC. Its assessment is characterised by simplicity and low effort. The mechanisms behind SARIFA suggest a tumour-promoting increased lipid metabolism and altered immune background, both showing new therapeutic avenues.

## Background

Worldwide, pancreatic cancer is the fourteenth most common malignancy but ranks seventh in cancer-related deaths [1] and is even prognosed to become the second most common cancer-related cause of mortality by 2030 [2]. The therapy still mainly relies on surgery (Whipple procedure) and adjuvant chemotherapy. However, in 85–90% of cases, tumours are primarily unresectable because of the infiltration of neighbouring structures or the presence of distant metastases [3]. Therapeutic improvements over the past two decades have been limited, and the disease is rightly described as a ‘difficult-to-treat’ entity with a five-year survival rate of only 11% [4]. Compared with other entities such as breast or lung cancer, there are only a few widely accepted prognostic factors routinely implemented in pathological diagnostic workups, including the factors of tumour-node-metastasis (TNM) classification, microsatellite instability status [5] and BRCA mutational analyses [6]; hence, there is a lack of further established and routinely applicable markers.

In our recent studies on gastric and colon adenocarcinomas, we established a new histomorphological biomarker called SARIFAs (stroma areactive invasion front areas), which proved to be of independent prognostic relevance in these entities. Also in prostate cancer, a prognostic value could be demonstrated [7]. By definition, a SARIFA is characterised as an area at the tumour invasion front where there is an absence of desmoplastic stroma reaction on malignant invasion in the surrounding inobtrusive tissue, hence leading to direct contact between tumour cells and adipocytes. Detectable on haematoxylin and eosin (H&E)–stained slides, without the necessity for additional immunohistochemistry, simple to learn, and assessable in a short period with low interobserver variability, SARIFAs can be easily implemented in routine diagnostic workflow [8, 9]. Moreover, SARIFA positivity reflects metabolic reprogramming in which tumour cells gain advantage from enhanced lipid supply.The important role of lipids in cancer became a major research line. In particular, lipid metabolism is a promising target for the development of new therapy concepts [10].

Our previous observations led to the question of whether SARIFAs also occur in pancreatic ductal adenocarcinoma (PDAC), an entity known for its pronounced stromal desmoplastic component, and if this concept could be adapted for a neoplasm with a considerably different biology compared with the originally addressed ones.

Therefore, we hypothesised that this phenomenon (i) also occurs in PDAC, (ii) is significantly prognostic and (iii) shows signs of an enhanced lipid metabolism. To confirm these hypotheses, we conducted the first analysis of a local PDAC patient collective and additionally explored the biochemical and immune background via immunohistochemistry.

## Methods

### Patient cohort and ethical approval

The study collection consisted of 174 patients who underwent the Whipple procedure at the University Medical Centre Augsburg between 2005 and 2015. The inclusion criteria were a postoperative survival of >30 days and histologically confirmed diagnosis of PDAC in the resection specimens. Histopathological diagnoses other than PDAC or death within the first 30 postoperative days led to exclusion. The sample size was not statistically determined prior to investigation.

Histologic subtyping was not investigated. Because of the limited number of cases, a division between test and validation collections, as recommended by REMARK [11] and STROBE [12] guidelines, could not be conducted. The study was performed in compliance with the Declaration of Helsinki. The protocol was evaluated and approved by the ethical committee of the Ludwig Maximilian University of Munich (reference: 22-0437), with no declaration of consent from the patients required.

The clinical data were derived from Tumour Data Management, University Hospital of Augsburg, and completed with the information acquired from the patient files. The endpoint of the study was overall survival (OS), which was measured from the moment of diagnosis to death of any cause or last registered follow-up (censored entries). The median follow-up was calculated using the reverse Kaplan–Meier method [13]. The estimated median follow-up for the whole study collection was 90 months (67–113) and did not differ significantly between SARIFA-positive and -negative cases.

### Histopathological SARIFA assessment

All given H&E-stained tumour slides (total 980, median 5 per case), each covering an area of approximately 220 mm^2^, were examined by two independent investigators (PG and BM) who were blinded to the clinicopathological data. A SARIFA was defined as the direct contact between at least five tumorous cells or a malignant gland and inconspicuous adipocytes at the invasion front, as described recently by our group [8, 9]. Representative images of both SARIFA-positive and SARIFA-negative cases are presented in Figure 1. Because the morphological feature of a SARIFA itself occurs at a high frequency in PDAC and not only at the invasion front, we decided to renounce the restriction of the invasion front and counted also intra-parenchymal interactions with adipocytes. Moreover, we established a quantitative cut-off for classifying a case as SARIFA positive. After evaluating an initial subset of 40 consecutive cases, a cut-off of SARIFA positivity in at least 66% of tumour slides revealed the highest prognostic potential. Following the independent assessment by two investigators, the cases with discrepant SARIFA scores were re-evaluated jointly by the same investigators, and a consensus diagnosis was made using a double-headed microscope.

**Figure 1.**
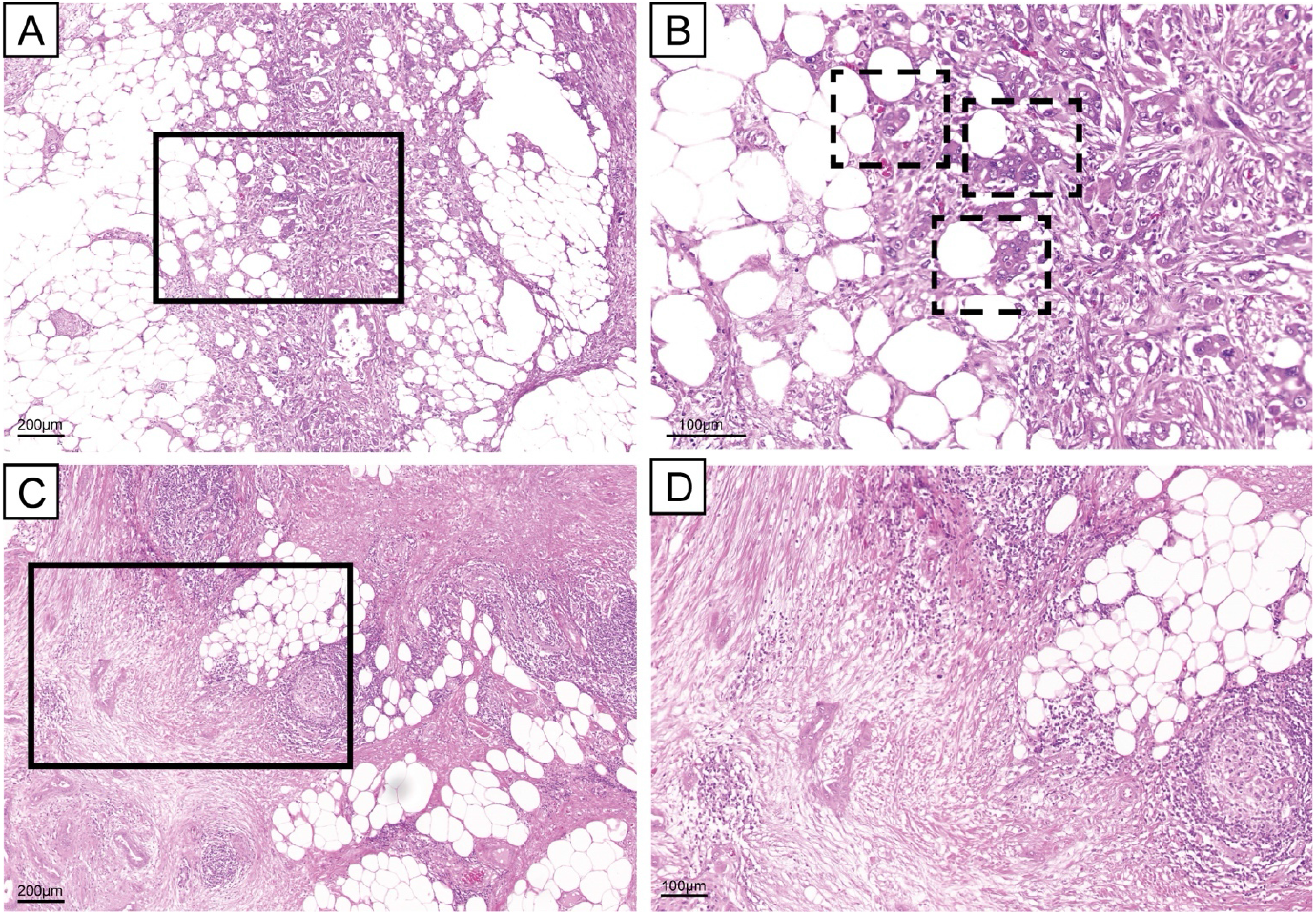
SARIFA-positive and -negative cases; H&E staining. A. Exemplary SARIFA-positive PDAC with tumorous cells directly adjacent to adipocytes at the invasion front; scale bar 200 µm. B. Detailed picture of SARIFAs (marked with a dashed line); scale bar 100 µm. C. SARIFA-negative PDAC with desmoplastic tumorous stroma separating malignant cells from surrounding fatty tissue; scale bar 200 µm. D. Detailed picture of SARIFA-negative PDAC; H&E; scale bar 100 µm. SARIFA – stroma areactive invasion front area; PDAC – pancreatic ductal adenocarcinoma.

### Immunohistochemical studies

Additional immunohistochemical staining was performed to analyse and compare the expression of fatty acid metabolism–related proteins and the role of macrophages in SARIFAs, here corresponding to the results of preceding analyses on gastric carcinoma [9]. FABP4, CD36 and CD68 immunohistochemistry was performed on 30 SARIFA-positive and 30 SARIFA-negative representative cases, using 2- to 4-µm-thick, whole-slide, formalin-fixed paraffin-embedded sections. The staining was performed on a Leica Bond RX automated staining system (Leica, Wetzlar, Germany) according to the automated immunohistochemical protocol optimised for use on this platform (antibodies and dilution in Supplementary Information Table S1). The assessment of FABP4 and CD36 at both the invasion front and tumour centre was conducted using the immunoreactive score, which is a seven-tier semiquantitative scoring system, as proposed by Remmele and Stegner [14]. Therefore, staining intensity and the percentage of positive tumour cells were evaluated to calculate the score accordingly. The number of CD68-positive macrophages was counted on a representative high-power field at the tumour centre and invasion front. Representative areas at the invasion front and tumour centre were selected by visual impression.

### Adipocyte morphometry

To verify and quantify a supposed delipidation of adipocytes, areas were digitally morphometrised. For that, H&E slides of 10 randomly selected SARIFA-positive and -negative cases from the above-described immunohistochemistry cohort were scanned using a 3D Histech Panoramic Scan II (3D Histech, Budapest, Hungary), and the morphometric measurements were performed using the CaseViewer 2.4 software (3DHistech, Budapest, Hungary). Two adipocytic areas each of the invasion front and of locations distanced from the tumour were analysed by one investigator (BM) by measuring the area of 4 to 13 adipocytes (mean: 10 ± 2) (Figure 2.).

**Figure 2.**
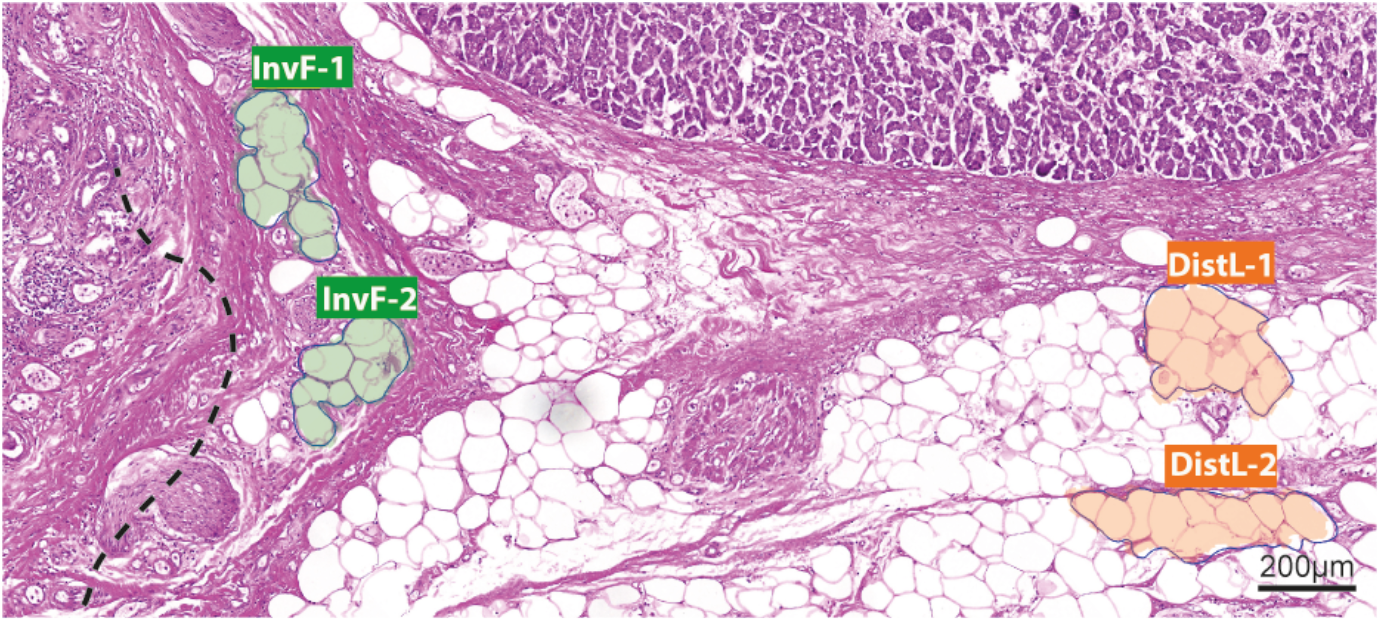
Principle of morphometric measurement of adipocytes. Exemplary PDAC slide with marked adipocytes at invasion front (InvF-1 and -2) and in distant locations (DistL-1 and -2), each with approx. 10 adipocytes; H&E; scale bar 200 µm. PDAC – pancreatic ductal adenocarcinoma.

### Statistical analysis

SPSS version 29.0 (SPSS, IBM, Chicago, IL, USA) and RStudio 2022.07.0 (R Foundation for Statistical Computing, Vienna, Austria) were used for statistical analysis. Correlations between clinicopathological data and SARIFA status were tested using Chi-squared tests or Fisher’s exact tests. The Kaplan–Meier method was used to depict the survival rates and the log-rank test to prove the significance of survival between the tested groups. The assessment of interobserver agreement was measured using kappa statistics. Relative risks were estimated by hazard ratios (HRs) calculated via Cox proportional hazard models.

Neither large language models nor artificial intelligence solutions were used in conducting the study.

## Results

### Clinicopathological characteristics

In the examined population of 174 PDACs, 21 patients were diagnosed with a pT1 tumour, 108 with a pT2 tumour and 37 with a pT3 tumour (according to the 8th Union for International Cancer Control staging system). In eight cases, the primary tumour (pT) category could not be ascertained due to a lack of precise information regarding tumour size in pathology reports. One hundred and thirty patients presented nodal and 102 distant metastases (eight during surgery, on suspicion of intraoperatively detected abdominal lesions). R-status was assessed according to the criteria proposed by Esposito et al. [15]. Both intrapancreatic and retroperitoneal resection margins were considered. A total of 120 (68.9%) patients received adjuvant chemotherapy (CTx) with different treatment regimens: Here, 78 were treated primarily with gemcitabine in monotherapy in a standard scheme of six courses and two with FOLFIRINOX schema. The remaining 40 patients received chemotherapy in other regimens (e.g., gemcitabine combined with erlotinib or radiotherapy) or did not complete the full treatment.

The median age at diagnosis was 68 years (range 42 to 85 years).

Overall, 54 cases (31%) were classified as SARIFA positive and 120 (69%) as SARIFA negative. SARIFA positivity was significantly associated with a higher rate of vascular invasion (p = 0.021) and lower frequency of adjuvant therapy (p = 0.006).

Other characteristics, including extension of pT, lymph node metastasis (pN) or distant metastasis, and R-status were not associated with SARIFA status (each p > 0.05). Detailed clinicopathological data are summarised in Table 1.

**Table 1.**
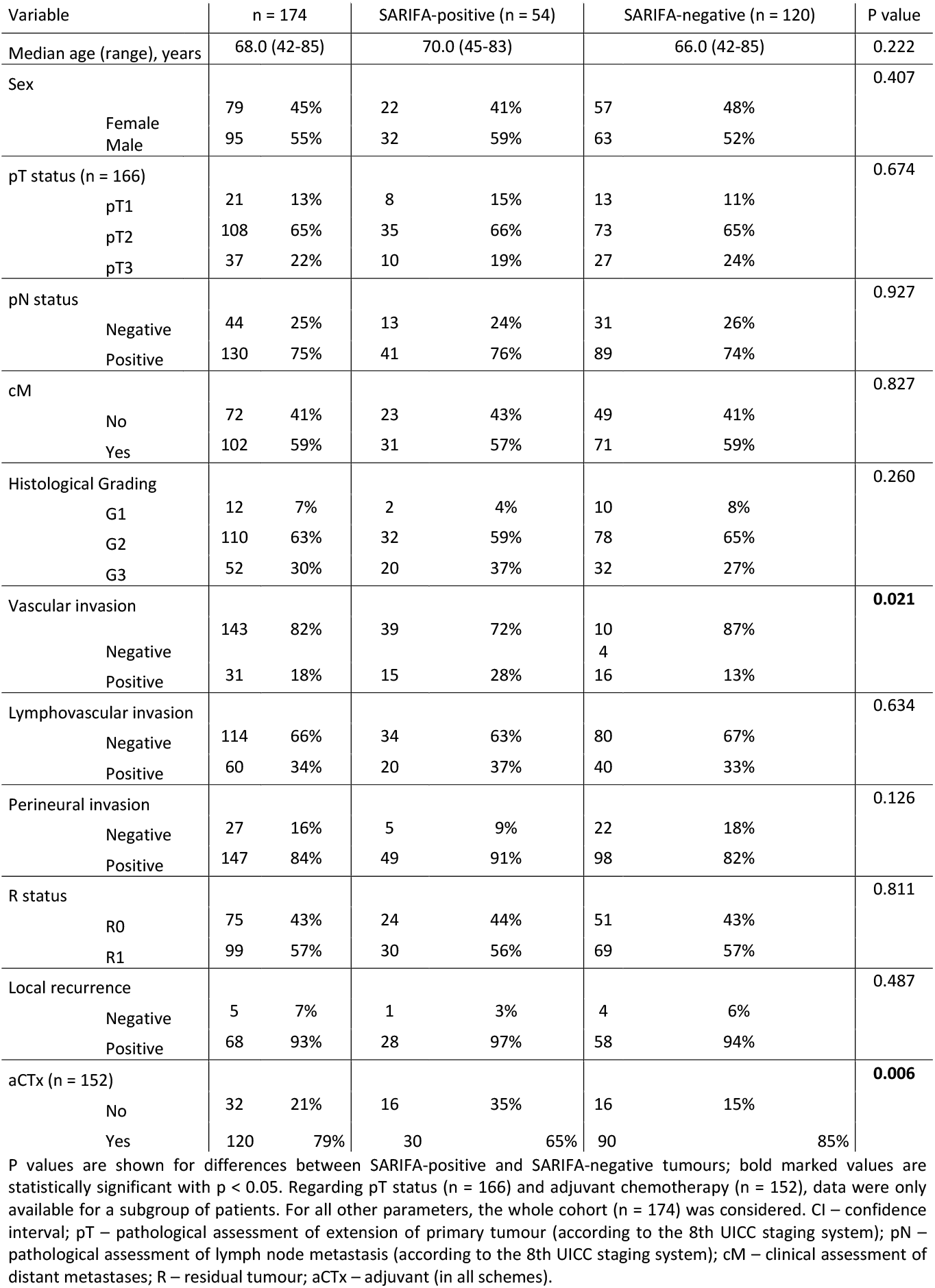
Clinicopathological characteristics.

Because obesity has previously shown significant correlations with the alternation of PDAC cell metabolism towards higher fatty acid uptake and a higher rate of tumour progression [16], we compared the body mass index between positive and negative patients (30 cases each) and found no significant correlation with the SARIFA status (p = 0.32; corresponding boxplot in Supplementary Information Figure S1).

### Interobserver variability

Considering distinctive stromal desmoplasia in PDAC, the assessment of SARIFA status appeared to be a demanding task. Nevertheless, the interobserver variability between the first and last author corresponded with a kappa value of 0.56, showing moderate interobserver agreement.

### Survival analysis

To analyse the prognostic relevance of SARIFA status in PDAC, we performed a Kaplan–Meier analysis and observed a distinct separation of survival curves (Figure 3., log-rank, p = 0.014). Our analyses showed that patients with SARIFA-positive PDAC had a significantly worse OS compared with SARIFA-negative cases (median OS: 9.9 months vs. 18.0 months, HR: 1.558, 95% CI 1.081–2.247, p = 0.018).

**Figure 3.**
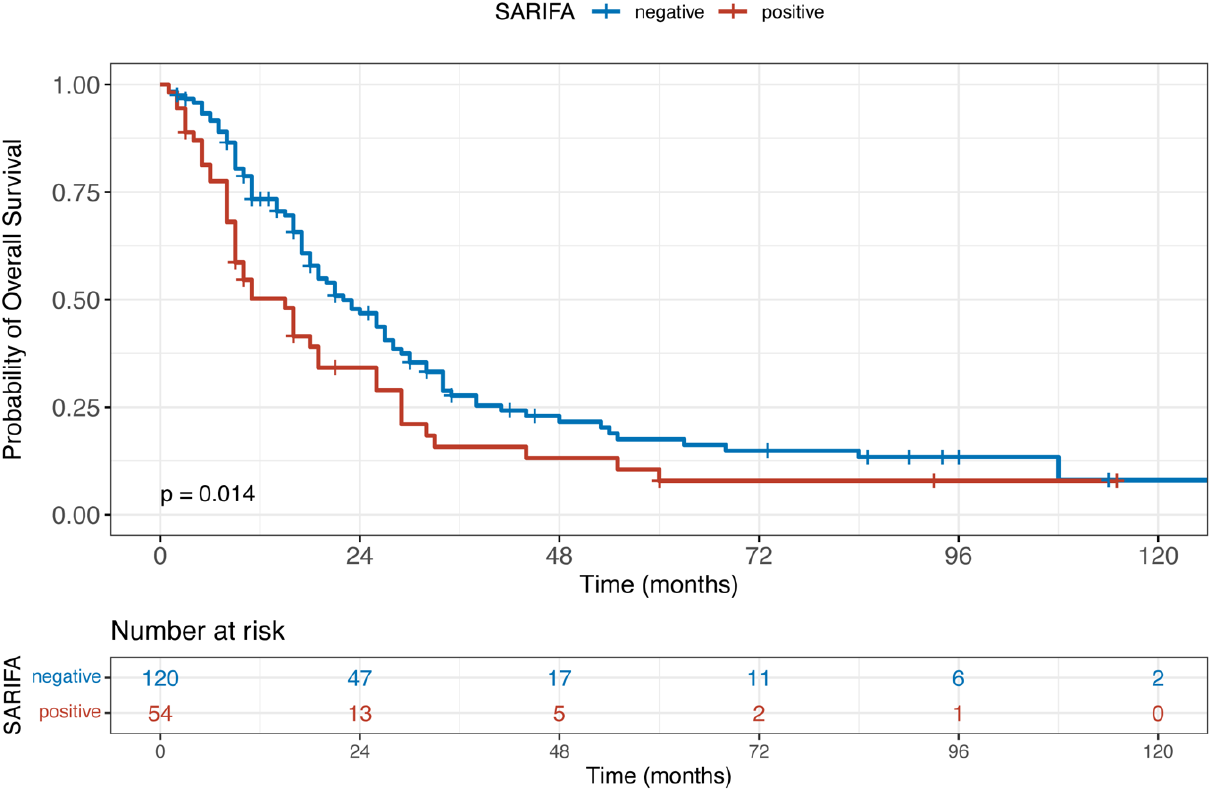
PDAC patient survival dependency on SARIFA status. Kaplan–Meier curve of patients with SARIFA-positive and SARIFA-negative PDAC. P value of the log-rank test. SARIFA – stroma areactive invasion front area; PDAC – pancreatic ductal adenocarcinoma.

To assess the prognostic relevance of SARIFA status compared with other risk factors, we performed uni- and multivariate Cox regression analyses. In the univariate analysis, patients’ age, tumour grading, adjuvant chemotherapy and SARIFA status were significantly related to worse OS (Table 2.). In the multivariate analysis, the following common risk factors were included: tumour grading, pT category, lymph node metastasis and invasion in blood or lymphatic vessels. Besides grading, SARIFA status remained significantly associated with shorter OS (p = 0.014), indicating that SARIFA status was an independent risk factor (Table 2).

**Table 2.**
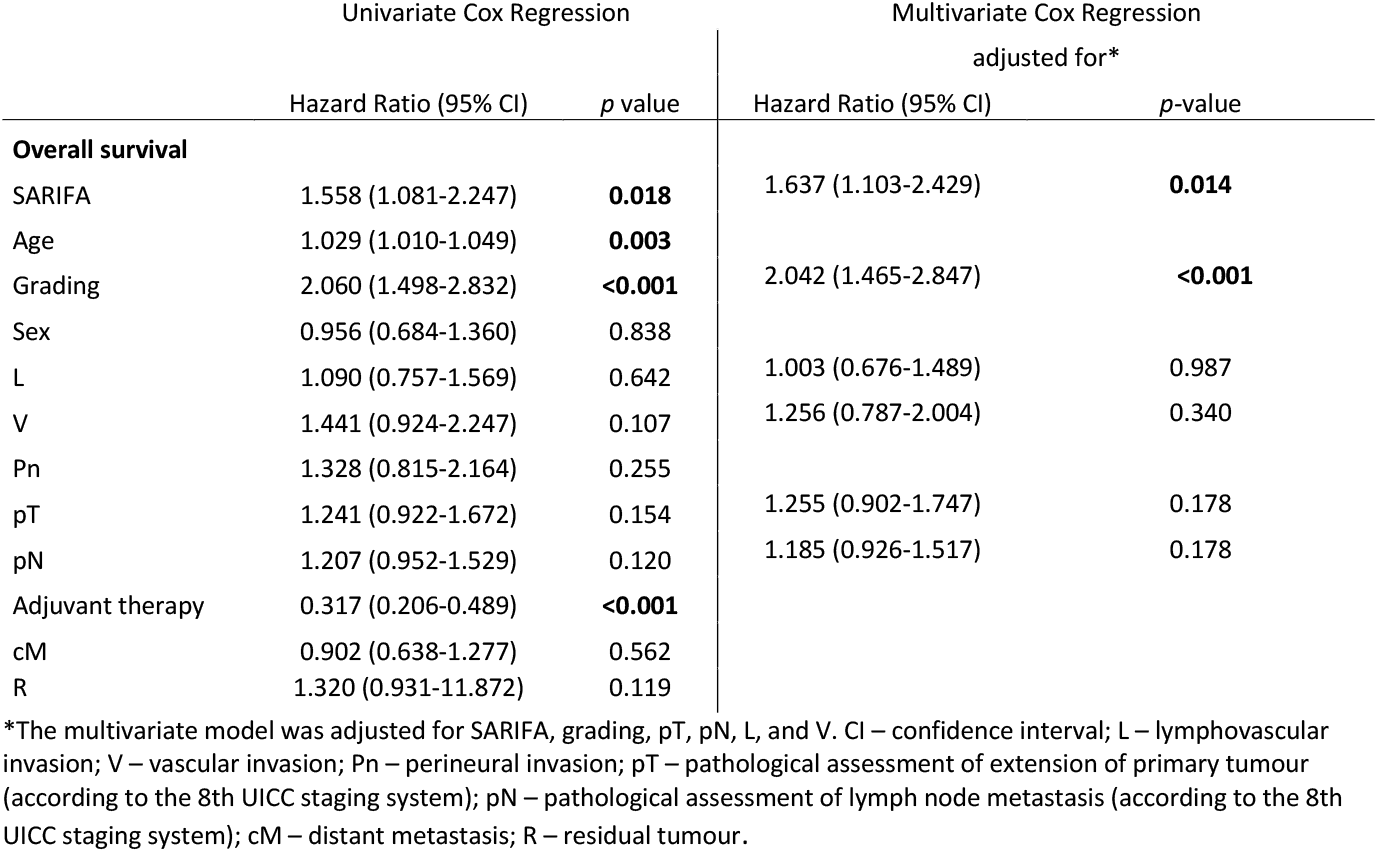
Uni- and multivariate Cox regression analysis regarding overall survival.

To assess the effect of SARIFA status on the impact of adjuvant therapy and, hence, whether SARIFA status may be predictive, we performed further subgroup analyses. Even though a higher percentage of patients with SARIFA-positive PDAC received adjuvant chemotherapy (16 cases, 35%), their OS was still lower than that of patients with a SARIFA-negative tumour, who were treated with adjuvant chemotherapy less frequently (16 cases, 15%). Within SARIFA-positive PDACs, adjuvant therapy was significantly associated with better OS (HR: 0.363, 95% CI 0.182–0.726, p = 0.004) but with a limited number of included patients (n total: 46, adjuvant therapy n = 30, no adjuvant therapy n = 16). This was also true within SARIFA-negative PDACs (n total: 106, adjuvant therapy n = 90, no adjuvant therapy n = 16) because adjuvant treatment was again associated with better OS (HR: 0.330, 95% CI 0.185–0.586, p < 0.001). These findings show that patients with PDAC benefit from adjuvant chemotherapy [17]. Corresponding Kaplan–Meier curves are provided in the Supplementary Information (Figure S2).

### Immunohistochemical expression of FABP4, CD36 and CD68 at SARIFAs

As mentioned above, we completed additional immunohistochemical studies focusing on lipid metabolism and tumour-associated macrophages at SARIFAs. Therefore, we investigated FABP4 and CD36 expression and the number of CD68^+^ macrophages. Tumour cells inSARIFA-positive cases showed higher expression of FABP4 at the invasion front than in SARIFA-negative cases (p < 0.0001). CD36 expression showed no statistically significant SARIFA-dependent changes (each p > 0.05). Moreover, CD68^+^ macrophages showed a higher density at the invasion front of SARIFA-positive than SARIFA-negative PDACs (p = 0.031). In SARIFA-negative regions, no differences regarding FABP4 and CD36 expression, as well as CD68^+^ macrophages, could be found (each p > 0.05). Immunohistochemical stains and the corresponding results are visualised in Figure 4.

**Figure 4.**
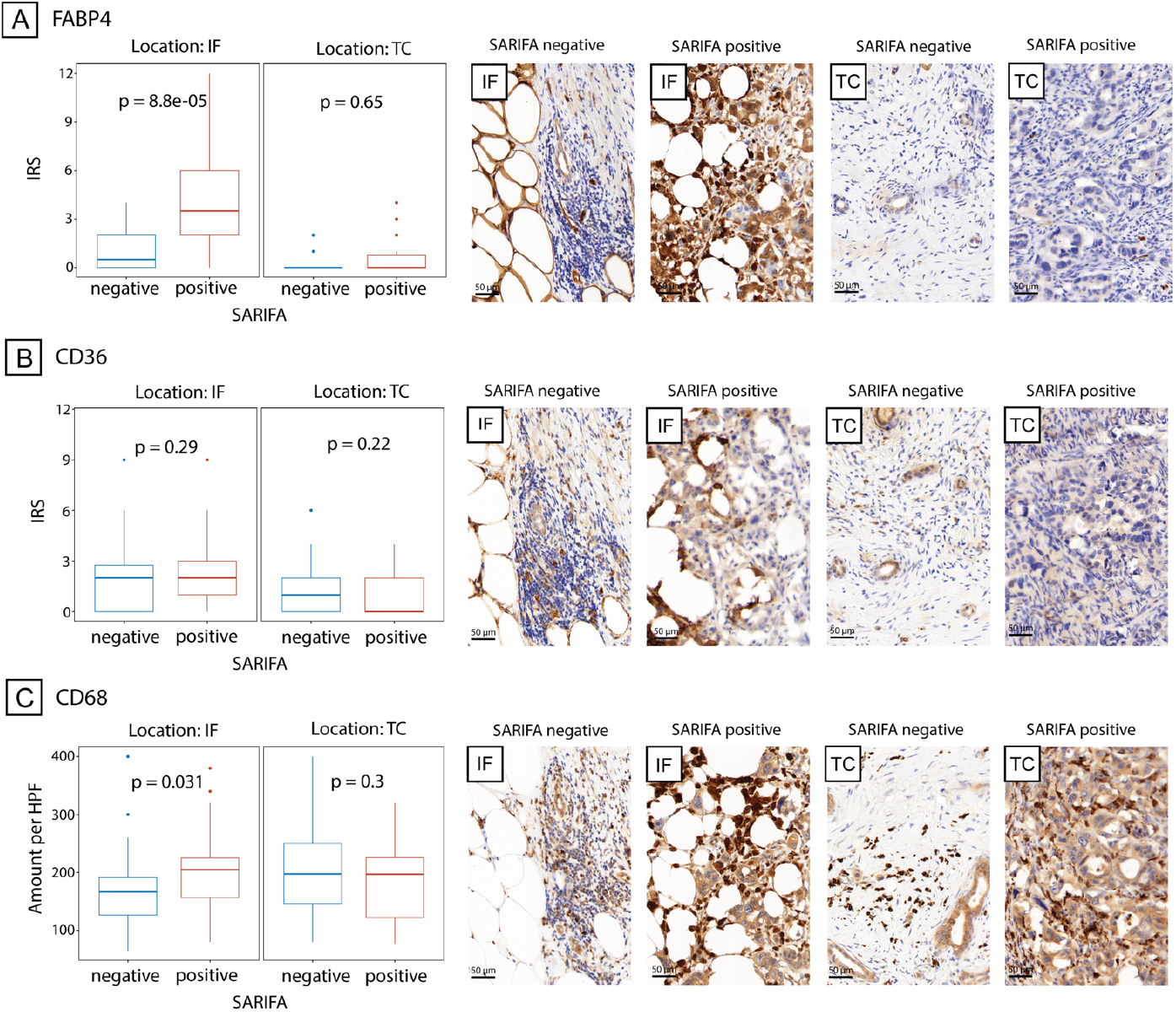
Expression of FABP4, CD36 and the presence of CD68-positive macrophages in SARIFA-positive and -negative cases at the tumour centre and invasion front. Boxplot showing differences in A. FABP4 expression, B. CD36 expression, and C. CD68+ macrophage count at invasion front (IF) and tumour centre (TC) with exemplary images. SARIFA – stroma areactive invasion front area; IRS – immunoreactive score; HPF – high-power field. Scale bar 50 µm.

### Adipocyte morphometry

The adipocyte areas in tumour-distanced locations did not differ between SARIFA-negative and -positive cases, with mean values of 5356 ± 1514 µm^2^ and 5140 ± 1559 µm^2^ (p = 0.659), respectively. The adipocyte areas at the invasion front were significantly smaller, with mean values of 4021 ± 1058 µm^2^ and 1812 ± 1008 µm^2^ for the SARIFA-positive and -negative cases, respectively. The area differences between the SARIFA-positive invasion front area and the other three parameters were highly significant (p < 0.001) (Figure 5.).

**Figure 5.**
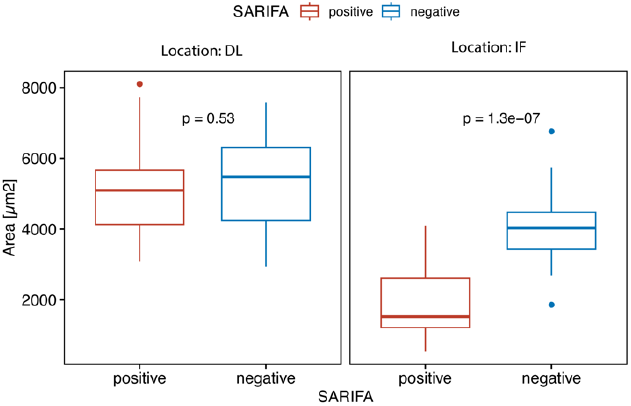
Adipocyte morphometry in SARIFA-positive and -negative cases at invasion front and locations distant from tumour. Boxplot showing differences in size of areas of approx. 10 adipocytes between SARIFA-positive and - negative cases in distant locations (DL) and invasion front (IF); SARIFA – stroma areactive invasion front area.

## Discussion

The role of lipid metabolism has gained increasing recognition. Lipid metabolism offers the potential of a new therapeutic target [10]. With SARIFAs, we recently introduced a new prognostic biomarker whose biological significance lies in detecting lipid-driven tumours. This effect has been demonstrated in gastric, colorectal and prostate cancer so far. In contrast to what might be expected, the occurrence of SARIFAs did not correlate with obesity [7, 9]. In the current study, we tested the hypotheses that SARIFA classification is applicable to and prognostic in PDAC and that there are signs of enhanced lipid metabolism.

Indeed, the SARIFA classification is applicable to PDAC. However, it had to be adapted considerably, especially to gastric and colorectal cancer. The restriction to evaluate only the invasion front had to be abandoned, and a quantitative cut-off had to be established. This meant that the evaluation became somewhat more challenging compared with the previous applications, resulting in a lower but still acceptable kappa value that is in the range of other histological features [18]. Moreover, ongoing research on the classification of PDAC using deep neural networks opens up perspectives for further improvement of the evaluation of SARIFAs and is reassuring regarding the reproducibility of assessment [19].

Compared with other tumour entities, in PDAC, there is a lack of biomarkers, and the 8th Union for International Cancer Control TNM staging system plays the most important yet debatable role in this context [20, 21], hence indicating the need for new biomarkers. Tumour budding, well established in colorectal cancer [22], did not enter routine and is, despite its broad acceptance as reliable prognostic biomarker, hampered by a relevant interobserver variability [23] because the standardisation and implementation of this marker has met some challenges, including demarcation of small-sized buds (up to five tumour cells) from surrounding cell-rich PDAC stroma, not only at the invasion front, but also at the tumour centre [24, 25]. DNA/RNA-based or subtype analysis and gene expression profiling are cost- and time-consuming assays and currently often have limited availability [5, 26-28].

In line with our findings, several studies deploying deep-learning algorithms on digitised slides of colorectal cancer patients were able to identify a morphologically similar phenomenon, one described as a ‘tumour adipose feature’ or ‘adipocytes close to tumour cells’, to be prognostically highly relevant [29-31]. These studies support our hypothesis regarding the high relevance of direct interactions between tumour cells and adipocytes. This is further strengthened by animal and in vitro PDAC models [32, 33], which have shown that adipocytes interact with and directly promote proliferation of malignant cells by increasing their fat uptake. To validate that this is also true in human tissue, we performed immunohistochemistry for CD36 and FABP4, two proteins that play major roles in lipid metabolism. Indeed, we were able to demonstrate prove significantly increased FABP4 expression, particularly at SARIFAs. The immunohistochemical expression of CD36, a multiligand translocase enabling transmembranous allocation of oxidised low-density lipoproteins, does not differ between SARIFA-positive and SARIFA-negative PDAC. These findings deviate from the analyses by Grosser et al. in gastric cancers, where CD36 was more strongly expressed in SARIFA-positive tumours [9], indicating that the regulation mechanism in these two entities differs. Therefore, the uptake of fatty acid could rely on an alternative transport mechanism like extracellular vesicles [34]. FABP4 is responsible for intracellular transportation and metabolism of fatty acids and was previously reported to be associated with poor prognosis in PDAC [35] and other malignancies [36], which is in line with our findings. Its upregulated expression in the context of direct contact between malignant cells and adipocytes, even in a highly glucose-dependent malignancy such as pancreatic cancer [37], suggests a more distinctive role of fatty acids as an energy source and supply of building blocks for cellular membranes in SARIFA-positive PDAC. The fact that adipocytes shrink when coming into contact particularly with tumour cells, as shown by our morphometric analyses, suggests adipocytes’ delipidation and uptake of lipids by the tumour cells. There is a large body of evidence indicating that lipids play a fundamental role in tumour progression[10, 38]. Metabolic reprogramming has been included in the hallmarks of cancers [39]. It seems likely that SARIFAs could serve as a biomarker that is not only prognostic but also effective for the selection of tumours that are particularly driven by lipids.

This could pave the way for new treatment approaches specifically targeting lipid metabolism in SARIFA-positive PDAC, for example, by using metformin, CPT1 or FABP4 inhibitors [10, 40-42] .

Among the several cell populations influencing both the growth and chemotherapy resistance of PDAC, tumour-associated macrophages drew our attention as an essential component of its microenvironment, playing a significant role in its biology [43, 44]. Moreover, CD68^+^ macrophages were upregulated at the SARIFAs in our study of gastric cancer [9]. In line with this, we observed higher concentrations of CD68^+^ macrophages at the invasion front of SARIFA-positive PDAC compared with SARIFA-negative cases, whereas in the tumour centre, there was no difference. Di Caro et al. showed that a higher density of macrophage infiltration at the tumour–stroma interface is associated with progression and distant metastasis of therapy-naïve PDAC [45] as a result of tumour-associated macrophages’ immunosuppressive activity. This mechanism could be co-responsible for both the development of SARIFAs and the non-favourable prognosis of SARIFA-positive PDAC cases, along with other alterations in local immune response [46].

The retrospective nature of the present study and the relatively low case numbers constitute its major limitations, with a need for further validation in prospective studies with possibly higher specimen amounts to conduct more precise stratification of our SARIFA effect. This is especially important because we could not establish a test and independent validation series, which is generally demanded for biomarker studies. The fact that we were previously able to prove the basic principle of SARIFAs in other entities several times might mitigate this limitation to some extent. The underlying biological mechanisms that lead to SARIFAs remain mostly unclear. Therefore, the mechanisms behind this morphological phenomenon, which seem to be of an immune nature [46], require further investigation, not only in PDAC but also in other entities showing our SARIFA phenomenon.

## Conclusions

The present study has confirmed the value of SARIFA status as a negative prognostic factor in PDAC. Compared with other novel biomarker approaches, which can only partly be evaluated on H&E-stained slides, SARIFA assessment is characterised by simplicity and low effort, enabling reliable patient stratification. The mechanisms behind SARIFAs suggest the major role of an increased tumour-promoting lipid metabolism and altered immune background. Therefore, based on our findings, we propose SARIFA status as a novel biomarker in PDAC that could not only help better stratify patients but also guide new therapeutic avenues by interfering in the lipid metabolism of tumour cells.

## Data Availability

All data produced in the present study are available upon reasonable request to the authors.

## List of abbreviations

CI: confidence interval
CTx: adjuvant chemotherapy
H&E: haematoxylin and eosin
HR: hazard ratio
OS: overall survival
PDAC: pancreatic ductal adenocarcinoma
pT: primary tumour
pN: lymph node metastasis
SARIFAs: stroma areactive invasion front areas
TNM classification: tumour-node-metastasis classification

## Declarations

### Ethics approval and consent to participate

Due to retrospective nature of the study no active intervention involving human participants and/or animals were applied. The study protocol was evaluated and approved by the ethical committee of the Ludwig Maximilian University of Munich (reference: 22-0437), with no declaration of consent from patients required.

### Availability of data and materials

The datasets generated throughout the analysis can be obtained from the corresponding author upon reasonable request.

### Authors’ contributions

B.M., P.G., N.G.R. and B.G. contributed to the study’s conception and design; P.G., N.G.R., B.G., F.S., A.P., J.W., G.S. and B.M. contributed to the data acquisition process; P.G. and N.G.R. contributed to the analysis and interpretation of the data. N.G.R. and B.M. contributed equally and share last authorship. All authors revised the article critically, contributed to it with reflective improvements and approved the final version. B.M. is the guarantor of this work and, as such, had full access to all of the data in the study and takes responsibility for the integrity of the data and the accuracy of the data analysis.

## Acknowledgements

We are grateful to Eva Sipos, Alexandra Martin and Christian Beul for their excellent technical assistance.

## Supplementary Information

**Fig. S1.**
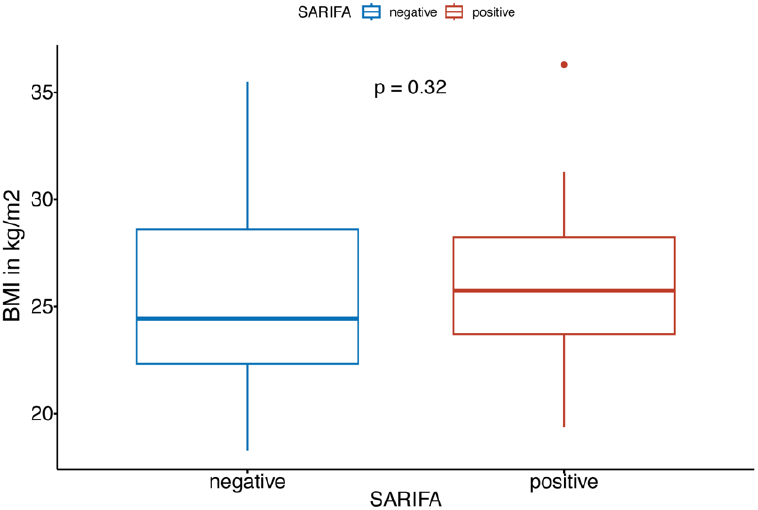
BMI differences between patients with SARIFA negative and positive PDAC. Boxplot showing differences in BMI between patients with SARIFA negative and positive PDAC; BMI – body mass index; SARIFA – stroma areactive invasion front area; PDAC – pancreatic ductal adenocarcinoma.

**Fig. S2.**
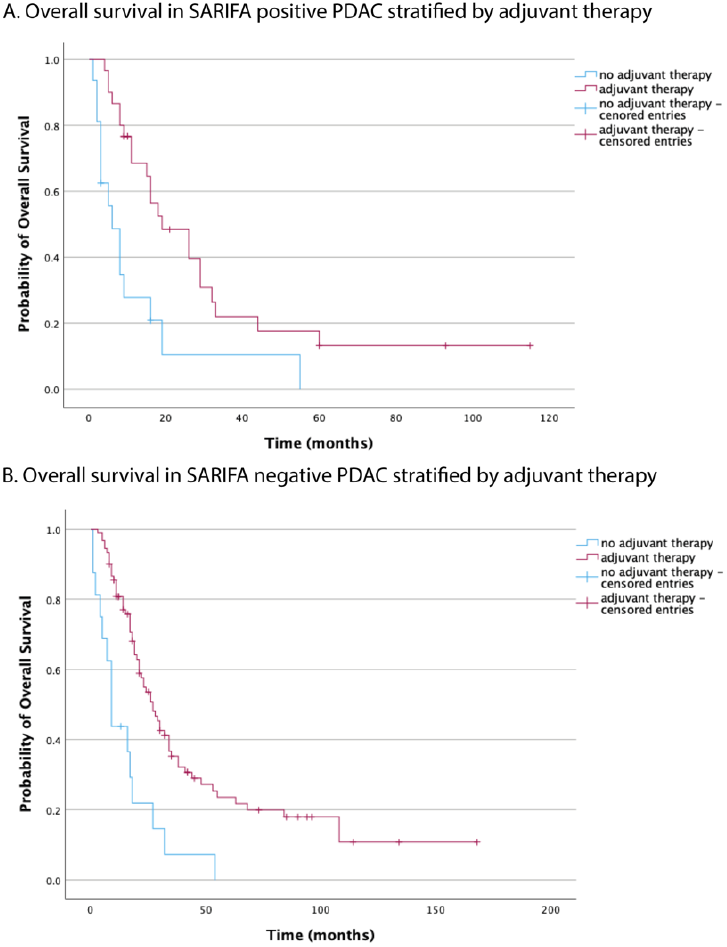
PDAC patient survival dependency on adjuvant therapy in SARIFA positive and negative groups. Kaplan–Meier curve of patients with SARIFA-positive and negative PDAC: survival dependency on adjuvant therapy; SARIFA – stroma areactive invasion front area; PDAC – pancreatic ductal adenocarcinoma.

**Table S1.** Antibodies and dilution used for immunohistochemical staining.

| Antigen | Clone | RRID | Supplier | Dilution | Positive control | Negative control |
| --- | --- | --- | --- | --- | --- | --- |
| CD36 | HPA002018 | AB_1078464 | Sigma Aldrich (St Louis, MO, USA) | 1:50 | Placenta | Internal negative control in the stained tissue and one slide per run omitting the primary antibody |
| CD68 | KP-1 | AB_1158192 | Cell Marque | 1:200 | Tonsil |  |
| FABP4 | ab13979 | AB_1951817 | Abcam | 1:500 | Liposarcoma |  |

